# The Role of Societal Aspects in the Formation of Official COVID-19 Reports: A Data-Driven Analysis

**DOI:** 10.1101/2020.10.28.20221572

**Authors:** Marcell Tamás Kurbucz, Attila Imre Katona, Zoltán Lantos, Zsolt Tibor Kosztyán

## Abstract

This paper investigates the role of socioeconomic considerations in the formation of official COVID-19 reports. To this end, we employ a dataset that contains 1,159 preprocessed indicators from the World Bank Group *GovData360* and *TCdata360* platforms and an additional 8 COVID-19 variables generated based on reports from 138 countries. During the analysis, a rank-correlation-based complex method is used to identify the time- and space-varying relations between pandemic variables and the main topics of World Bank Group platforms. The results not only draw attention to the importance of factors such as air traffic, tourism, and corruption in report formation but also support further discipline-specific research by mapping and monitoring a wide range of such relationships. To this end, an R Notebook is attached that allows for the customization of the analysis and provides up-to-date results.

## Introduction

Research on the COVID-19 pandemic has grown rapidly since the outbreak of the disease; however, despite the enormous media attention on countries’ reports, only a few articles address the number of officially reported cases and deaths as a social phenomenon. As many studies have pointed out, there is a significant discrepancy between the officially confirmed data and recently published estimates (see, e.g., [1, 2]). However, what do these data reflect, beyond the true nature of the virus? Of the few articles dealing with this question, [3] examined the protective effect of BCG on COVID-19 infections and the death toll while using indicators such as the Human Development Index (HDI), per-capita GDP, and urban population percentage as additional control variables. Moreover, in line with [4], they applied the Corruption Perception Index (CPI) as a proxy for the reliability of reported COVID-19 data. [5] showed that more democratic political institutions experienced deaths on a larger per-capita scale and sooner than did less democratic countries, and based on [6], the population size and government health expenditure are strongly related to COVID-19 cases.

In contrast to these (mostly) discipline-specific studies, our goal is to map, analyze, and monitor a wide range of such relationships in time and space by applying a data-driven approach. In this paper, we employ a rank-correlation-based complex method and a dataset that contains 1,159 preprocessed indicators from the World Bank Group *GovData360* and *TCdata360* platforms and an additional 8 COVID-19 variables. The results not only draw attention to the importance of factors such as air traffic, tourism, and corruption in report formation but also support decision makers and discipline-specific research by providing an R Notebook that allows for the customization of the analysis and provides up-to-date results.

The paper is organized as follows. The next section introduces the data and methodology used during the calculations. Then, the results are presented and discussed. Finally, the last section provides the conclusions and proposes future research directions.

## Data and methodology

### Joint dataset of GovData360, TCdata360 and COVID-19 reports

This paper follows the steps of [7] when creating a linked database of governance, trade, and competitiveness indicators with COVID-19 reports. (To derive the up-to-date dataset, we use the author’s R source code, which is publicly available at [8].) Former indicators were obtained from the *GovData360* and *TCdata360* platforms using the *data360r* (version: 1.0.8) R package [9]. From these platforms, only annual indicators from 2015 and later were collected, and their missing values were replaced with previous annual values in descending order by year until 2015. During preprocessing, indicators (columns) where the ratio of missing values exceeded 50% were filtered out. Then, the same filtration was applied above 25% in the case of countries (rows). Finally, highly correlated variables and variables with near-zero variances were removed, and the standardized form of the retained 1,159 indicators was connected with 8 COVID-19 variables, generated based on the official reports of 138 countries [10]. Note that auxiliary indicators measuring the number of data sources and standard error were also filtered out, and variables with near-zero variances were eliminated using the default settings of *nearZeroVar* function contained by the *caret* (version: 6.0-85) R package [11]. The presented data were compiled on July 22, 2020. Table 1 shows the description of the variables in the structure of the final dataset.

**Table 1.**
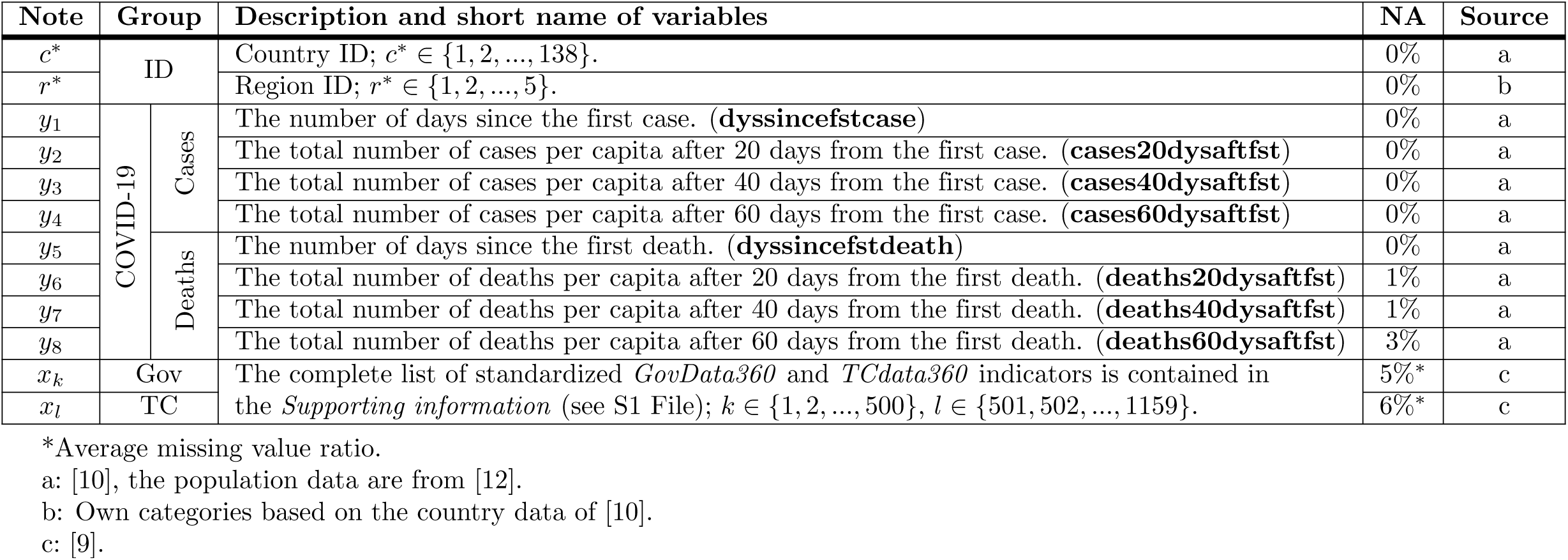
Variable description.

### Community-based model reduction

Our goal is to map the time- and space-varying relationship between COVID-19 (furthermore dependent) variables (*𝒴* := {*y*_1_, ‥, *y*_*M*_}) and indicators from *GovData360* and *TCdata360* platforms (furthermore independent variables, 𝒳:= { *x*_1_, ‥, *x*_*N*_}). To obtain an easily interpretable, comprehensive picture from these connections, similar *GovData360* and *TCdata360* indicators are grouped and characterized by latent variables. The applied steps are as follows.

First, standardized independent variables that have higher absolute (Spearman) rank correlation than an *α* parameter with at least one dependent variable are selected and denoted as *X* ⊆ *𝒳*. Formally:

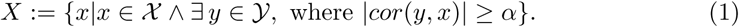

Then, the rank correlation matrix of the selected variables is used as an adjacency matrix **A**, in which absolute rank correlation values below a *β* parameter are substituted by 0. Formally:

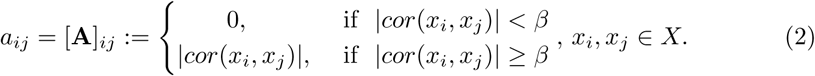

Note that the adjacency matrix **A** defines a network, where the vertices are the selected variables (*V* = *X*), edges are indicated by the nonzero values (*e*_*ij*_ ∈ *E* ⇔ [**A**]_*ij*_ = *a*_*ij*_ > 0) and their weight is the absolute rank correlation between the selected variables (*w* : *E* → ℝ^+^, *w*(*i, j*) = *a*_*ij*_). (Note that the same strategy was applied by, e.g., [13] and [14], to visualize variable similarity.)

To group similar variables, our goal is to separate this network into groups of vertices that have fewer connections between them than inside the communities. In the literature, this task [15] is referred to as modularity-based community analysis (see, e.g., [16]) or simply community detection (see, e.g., [17]). Although the proposed method may seem more complicated than traditional model reduction methods, because of the large number of variables and relatively few observations, they cannot be used. In addition, the visualization of the (correlation) network facilitates control over community formation (especially if *N* is large). This benefit is realized by using the Louvain community detection method [18] with an associated filtrating procedure that gains heterogeneity between the groups by eliminating weakly connected group members.

After Louvain community detection, we obtain *C* := {*c*_1_, ‥, *c*_*n*_} communities on *G*(*V, E*), which specifies 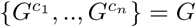 partitions of network *G*. As a next step, each community is represented by a single composite (so-called latent) variable 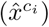 obtained by the weighted linear combination of member variables:

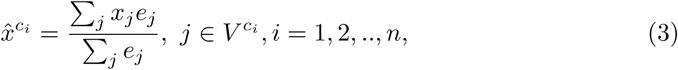

where

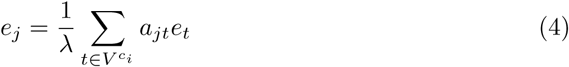

is the eigenvector centrality of node *j*, and *λ* > 0 is a constant. Louvain modularity and eigenvector centrality were calculated using the *cluster louvain* and *eigen centrality* functions of *igraph* (version: 1.2.4.2) R package, respectively [19]. Note that the use of eigenvector centrality as a weight ensures that deeper embedded variables (within the given community) play a greater role in the formation of the latent variable. (Also note that the use of standardized independent variables results in standardized latent variables). To increase the homogeneity of communities, we calculate the absolute (Spearman) rank correlation of each variable within the community *i* with the related latent variable 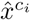, and variables that have weaker absolute rank correlation than a *γ* parameter are removed from their communities. Finally, the steps of this paragraph (from community detection to filtration) are repeated until no more variables can be eliminated.

At the end of the process, we rank communities (characterized by latent variables) by their absolute rank correlation with dependent variables. Then, we select the top *𝒞* ≤ *n* interpretable communities and investigate their relationship with the dependent variables through their absolute rank correlation coefficients. To examine the regional differences in addition to the study of time-varying relationships, these correlations are identified as well on the subset of different regions (see variable *r*^*\**^ in Table 1). The calculation steps are summarized in Fig 1.

**Fig 1.**
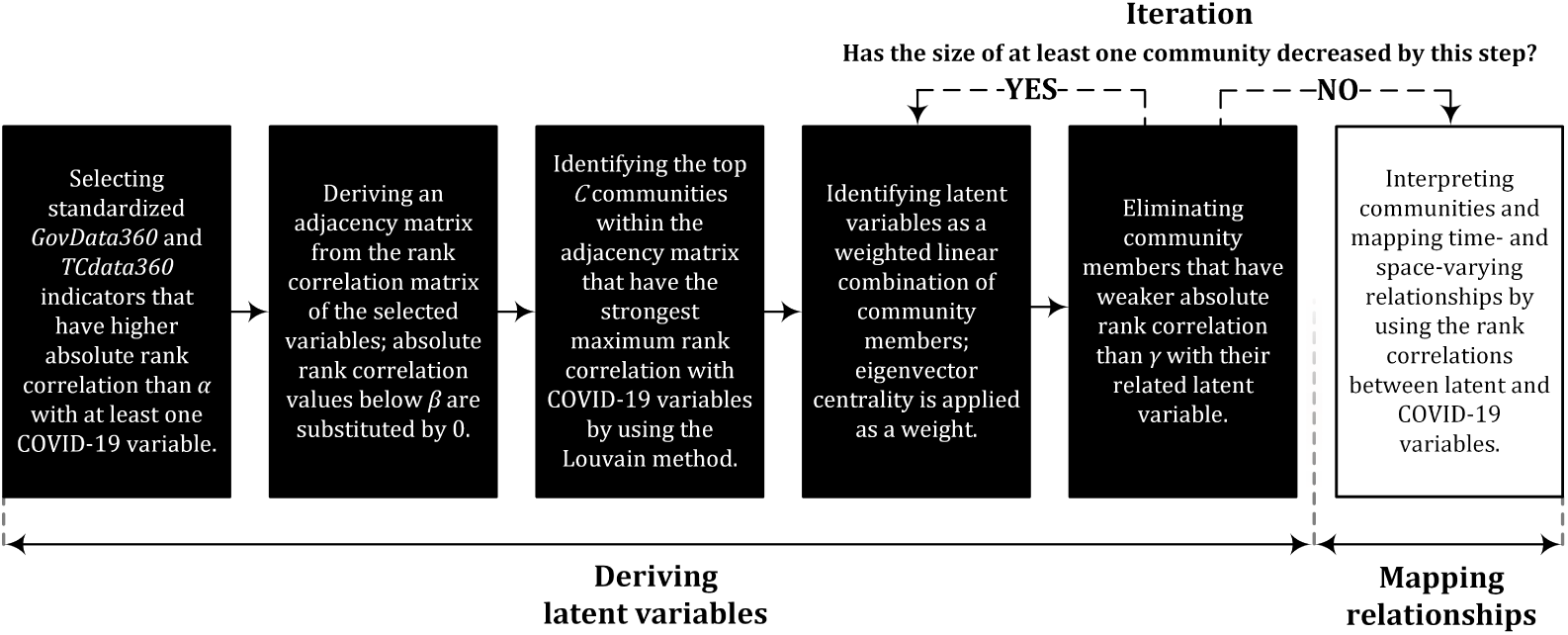
Calculation steps.

## Results and discussion

Following the calculation order presented above, we first illustrate and interpret the results of community detection to identify the most important topics reflected in official COVID-19 reports. Then, we investigate the time- and space-varying relations of these communities with different pandemic variables.

### Topics most related to COVID-19 reports

When setting *α, β*, and *γ* parameters, our goal was to group the widest possible range of important *GovData360* and *TCdata360* indicators without obtaining communities that are difficult to interpret. To accomplish this goal, we set the parameters as *α* = 0.535, *β* = 0.828 and *γ* = 0.770, which resulted in a network containing 319 indicators (vertices) and 1,669 edges, representing the strong correlations among the indicators. Fig 2 illustrates five communities (𝒞 = 5) detected within this network and helps their interpretation with word clouds generated based on the names of member indicators. Word clouds were constructed after text cleaning and preprocessing by using the *wordcloud* (version: 2.6) R package [20].

**Fig 2.**
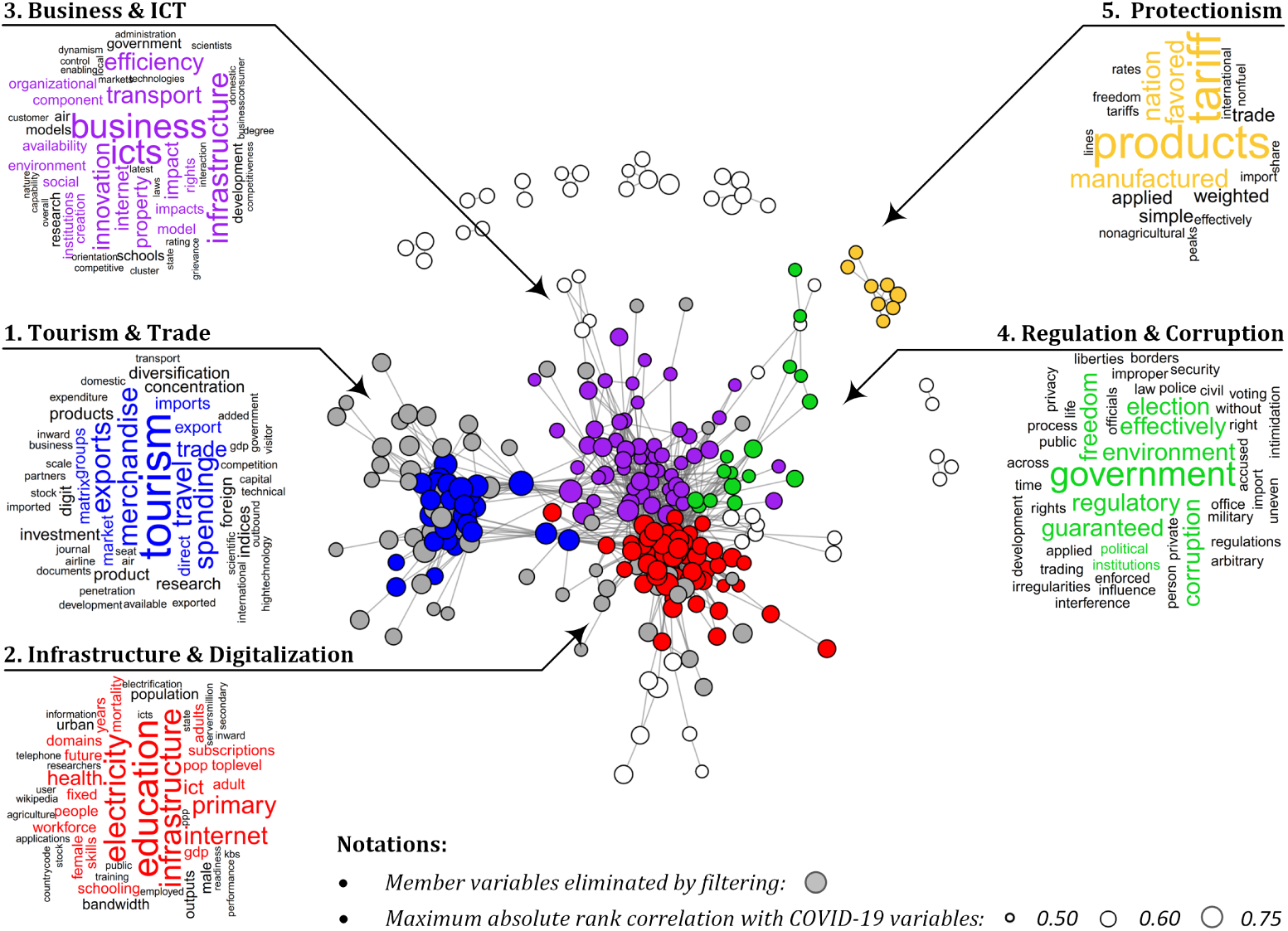
Detected communities.

As Fig 2 shows, the topics most related to official COVID-19 reports are (1) *tourism and trade*, (2) *infrastructure and digitalization*, (3) *business and ICT*, (4) *regulation and corruption*, and (5) *protectionism*. From these, *tourism and trade* associates with the flow of people and goods, as reflected by the most frequent terms such as *tourism, travel, merchandise*, and *imports*. The contribution of the database also confirms this finding since most indicators of this community are part of the *World Travel & Tourism Council* (27%), *United Nations Conference on Trade and Development Statistics* (19%), and *World Integrated Trade Solution* (15%) datasets. The second community describes the infrastructure, especially in the field of digitalization, including variables such as *ICT access, public services* and *secure internet servers/million pop*. Most of these variables are derived from the *World Economic Forum Global Competitiveness Index* (34%), *Global Innovation Index* (24%), and *World Development Indicators* (17%) databases. The third community, so-called *business and ICT*, is adjacent to *infrastructure and digitalization*. As its name suggests, it is in connection with information and communication technology (ICT); however, this community focuses more on business aspects such as *innovation, efficiency*, and *competitiveness*. The group *regulation and corruption* includes variables such as *regulatory quality, political environment*, and *corruption*. The sources of most of these variables are the *World Justice Project - Rule of Law* (31%), Global Innovation Index (25%), and Global State of Democracy (19%). Finally, the fifth community is labeled *protectionism* because all of its variables are related to *tariffs*. The variables for each community are detailed in the *Supporting information* (see S1 File).

### Relations with COVID-19 reports

To visualize the absolute rank correlation between the COVID-19 variables and the communities characterized by latent variables, radar charts are employed. In Fig 3, these relations are classified into three groups. The first focuses on the time elapsed since the first registered data, while the other two relate to the officially reported cases and deaths per capita aggregated by using different time windows.

**Fig 3.**
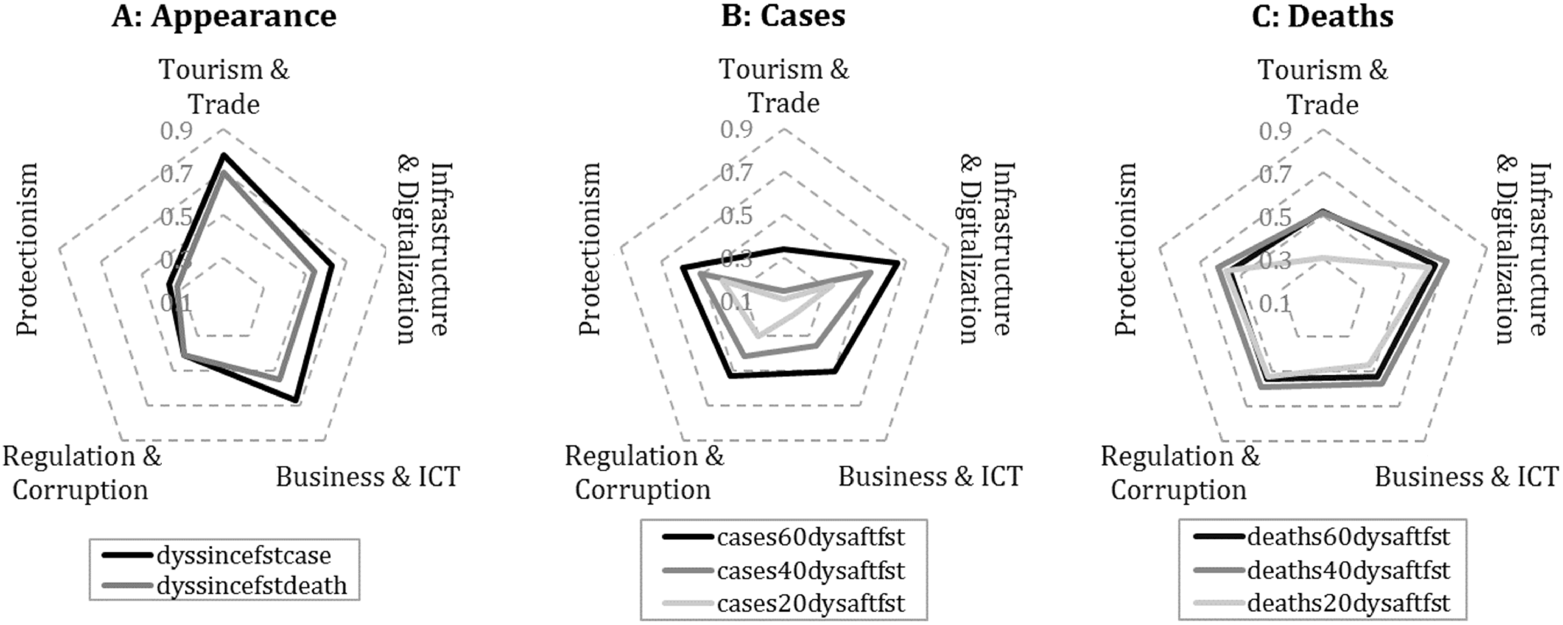
Absolute rank correlation between COVID-19 and latent variables.

As Fig 3 shows, indicators measuring the appearance of the virus are strongly correlated with tourism- and trade-related activities. Taking a closer look at the standalone variables within this community, *GCI 4*.*0: Air transport* and *outbound travel and tourism expenditure* have the strongest (Spearman) rank correlation coefficients with the days passed since the first case was reported (0.789 and 0.777, respectively). Moreover, this COVID-19 indicator has a strong connection with variables of international trade as well, such as *Merchandise: Trade matrix by product groups, imports* (0.755) from the same community. In light of these close relationships, it may be surprising to find that this community has a relatively weak connection with the reported number of cases and deaths; however, increased controls at airports and the rapid closure of borders could be reflected in this result.

In contrast to *tourism and trade*, these COVID-19 indicators are closely linked to the other four communities, especially to *infrastructure and digitalization*. From this community, variables related to digital development such as *fixed broadband subscriptions (per 100 people)* and *online creativity* show the strongest positive (Spearman) rank correlation with the number of deaths per capita (0.659 and 0.653, using the 60-day time window), which suggests that a significantly higher death toll has been reported by more developed countries. It is also reflected by the positive correlation of this COVID-19 variable with *A. Health* indicator calculated from healthy life expectancy (0.582) as well as by its strong negative relationship with *GCI 4*.*0: Exposure to unsafe drinking water* (− 0.676).

Based on these results, while the data suggesting the appearance of the virus seem to be reliable and relatively easy to explain, reports on cases and deaths appear highly distorted. On the one hand, this distortion may be a consequence of the poor health infrastructure that makes measurement difficult, but on the other hand, political interests could also be tied to underreporting. Since the *regulation and corruption* community’s *regulatory quality* and *freedom from corruption score* indicators have a strong positive correlation with the reported number of cases (0.560 and 0.533, respectively) and deaths (0.565 and 0.523, respectively, using the 60-day time window), the reports of countries with higher levels of corruption seem much less authentic. Furthermore, detected communities contain strikingly many indicators related to the development of the information society, which counteracts disinformation.

To support discipline-specific research, we detailed the correlations of each member variable with different COVID-19 indicators in the *Supporting information* (see S1-S5 Tables). Similar to latent variables (see Fig 3), these standalone indicators typically show increasing correlations with time window expansion; however, this change over time can vary significantly from region to region.

### Regional differences

To examine how the results presented in the previous subsection differ from region to region, countries are divided into four groups by using the *region ID* variable (denoted as *r*^*\**^ in Table 1). These groups are *Europe, Asia, Americas, Africa*, and *Oceania*; however, the last group was omitted from the investigation due its small sample size (2 countries). The regional differences in the relations of COVID-19 and latent variables are presented in Fig 4.

**Fig 4.**
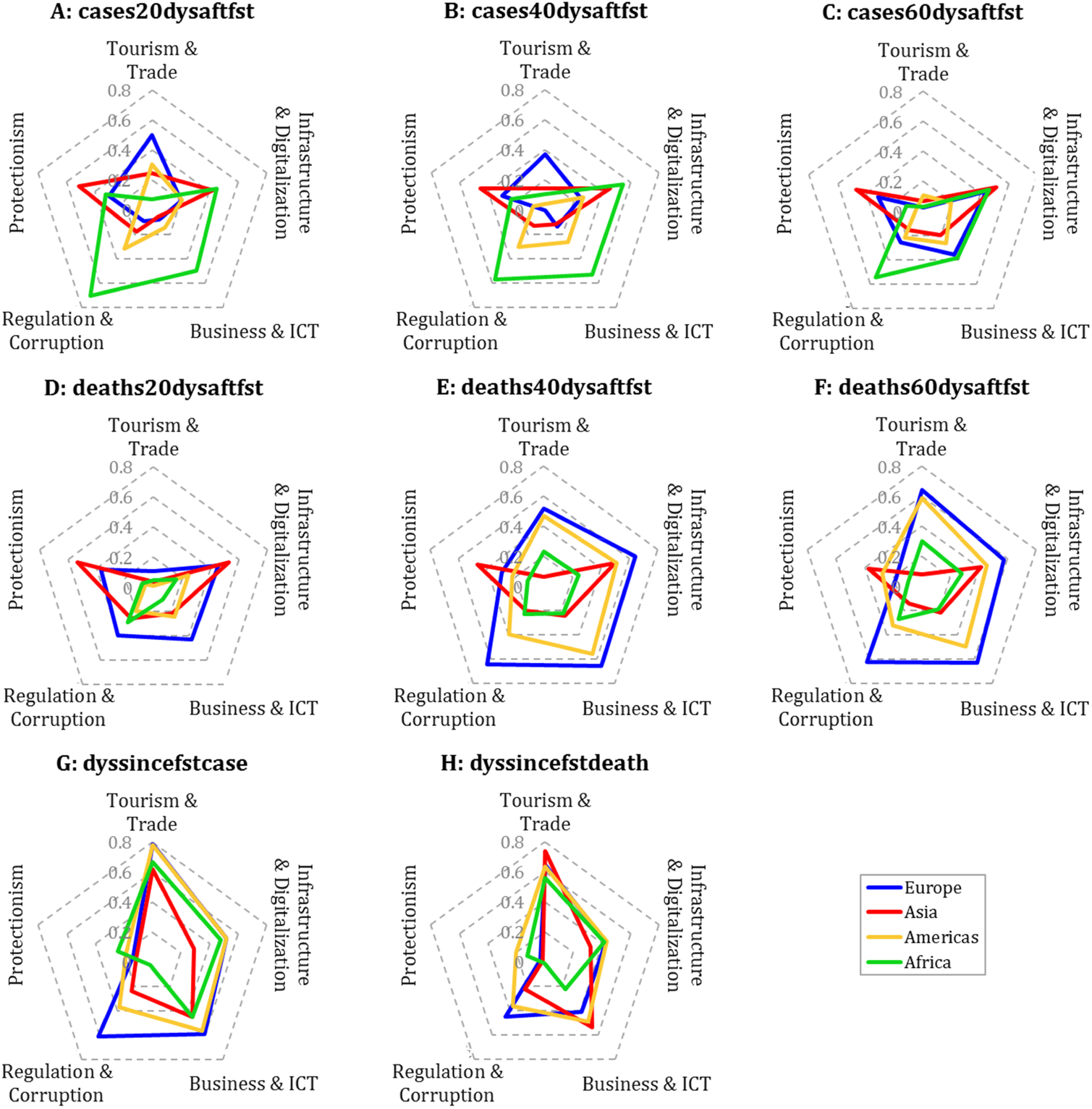
Absolute rank correlation between COVID-19 and latent variables by region.

Based on Fig 4, we can conclude that the impact of *tourism and trade* on the spread of the virus is significant regardless of region; however, the variable measuring the appearance of the first case shows the highest (Spearman) rank correlation with this community in the *Americas* and *Europe*. In these two regions, variables such as *GCI 4*.*0: Air transport* (0.803 and 0.791, respectively) and *government spending on travel and tourism service* (0.881 and 0.715, respectively) have one of the highest correlations with days elapsed since the first case. Moreover, in the *Americas* and *Europe*, this community, and especially its tourism- and air-transport-related indicators, shows an increasingly close relationship with the number of registered deaths per capita as the time window expands. Accordingly, regulations on foreign travel restrictions and airport controls are particularly important in these regions.

Next to the *Americas* and *Europe*, in *Asia*, variables measuring the spread of the virus are also strongly tied to the *tourism and trade* community, but these variables have a stronger rank correlation with the data related to first death. Unlike other regions, reports from *Asian* countries are mostly related to *infrastructure and digitalization* and *protectionism* communities; however, even these relations appear weak in comparison with the relationships detected in other regions. To obtain stronger ties, it may be worthwhile to map the topics that contain the most important variables separately for this region.

Finally, reported data both in *Africa* and *Europe* have remarkably close connections with the *regulation and corruption* community, especially with indicators such as *political environment* and *freedom from corruption score*. While in *Africa*, these variables are typically related to reported case numbers (0.568 and 0.551), in *Europe*, they show a stronger correlation with deaths (0.495 and 0.583, using the 60-day time window, respectively), which suggests that the reports of these regions are less credible. Note that the correlations of standalone variables calculated on different regional subsamples are contained in the *Supporting information* (see S2 File).

## Conclusions and future work

Although some of the recent studies have already investigated the relationship of COVID-19 data with different socioeconomic indicators, the role of societal considerations in the formation of official COVID-19 data is not yet clear. In contrast to these studies, our goal was to map, analyze, and monitor a wide range of such relationships in time and space by applying a data-driven approach. To this end, we employed a rank-correlation-based complex method and a dataset that contains 1,159 preprocessed indicators from the World Bank Group *GovData360* and *TCdata360* platforms and an additional 8 COVID-19 variables generated based on the officially reported number of cases and deaths.

From our results, the topics most related to official COVID-19 reports are *tourism and trade, infrastructure and digitalization, business and ICT, regulation and corruption*, and *protectionism*. By examining these topics and the variables they compress, we found that tourism- and air-transport-related variables are key factors in the spread of the virus, especially in the *Americas* and *Europe*. In these two regions, the variables of the *tourism and trade* community show close connections with the reported death toll as well, which also emphasizes the importance of regulations on foreign travel restrictions and airport controls. In addition, the number of reported cases and deaths seems unreliable since developed countries generally reported more cases and deaths than developing countries. In line with the results, the two possible reasons for underreporting may be the poor health infrastructure that makes measurement difficult and the political will that is opposed to exploring and presenting the real epidemiological situation. Accordingly, we experienced the closest relationship between the level of corruption and reported data in *Eurpoe* and *Africa*.

Using the proposed analysis, further interesting regional and temporal patterns can be identified, as the data will be updated over time. To support this research, we attach an R Notebook file (see S3 File) that not only updates the dataset but is also able to conduct all the analysis steps, including variable filtering and the compilation of figures. As a further advantage, this source code can be easily customized and allows researchers to apply arbitrary time frames during the analysis. Finally, in the *Supporting information*, we provide all the relationships identified during the analysis to support discipline-specific investigations.

## Data Availability

The datasets are available in the Supplementary Material.

## Acknowledgments

Supported by the ÚNKP-20-3 New National Excellence Program of the Ministry for Innovation and Technology and by the Research Centre at Faculty of Business and Economics (No. PE-GTK-GSKK A095000000-1) of University of Pannonia (Veszprém, Hungary).

## Supporting information

**S1 Table.** Standalone correlations in the *tourism & trade* community. Spear-man rank correlations between COVID-19 variables and indicators of the *tourism & trade* community in the worldwide dataset.

| Name | Type | dyssincefstcase | cases20dysaftfst | cases40dysaftfst | cases60dysaftfst | dyssincefstdeath | deaths20dysaftfst | deaths40dysaftfst | deaths60dysaftfst |
| --- | --- | --- | --- | --- | --- | --- | --- | --- | --- |
| Trade, competition, & market scale | rank* | 0.69 | -0.15 | 0.07 | 0.28 | 0.68 | 0.19 | 0.42 | 0.43 |
| Scientific and technical journal articles | value | 0.77 | -0.19 | 0.07 | 0.25 | 0.69 | 0.20 | 0.43 | 0.44 |
| Travel and Tourism direct contribution to GDP | usd nominal | 0.75 | -0.28 | -0.04 | 0.15 | 0.66 | 0.08 | 0.32 | 0.35 |
| Outbound Travel & Tourism Expenditure | usd nominal | 0.78 | -0.19 | 0.04 | 0.25 | 0.70 | 0.18 | 0.41 | 0.44 |
| Index Of Export Market Penetration | value | 0.74 | -0.19 | 0.06 | 0.27 | 0.71 | 0.30 | 0.51 | 0.53 |
| Merchandise: Trade matrix by product groups, imports | value | 0.76 | -0.27 | -0.01 | 0.20 | 0.67 | 0.16 | 0.40 | 0.41 |
| Government spending on travel and Tourism service | usd nominal | 0.67 | -0.18 | 0.05 | 0.23 | 0.68 | 0.17 | 0.41 | 0.42 |
| Capital investment in Travel and Tourism | usd real | 0.71 | -0.21 | -0.03 | 0.15 | 0.62 | 0.09 | 0.32 | 0.35 |
| Services, etc., value added (current US\$) | value | 0.70 | -0.27 | -0.02 | 0.17 | 0.66 | 0.08 | 0.33 | 0.36 |
| Business Tourism Spending | usd nominal | 0.74 | -0.29 | -0.06 | 0.15 | 0.64 | 0.08 | 0.32 | 0.37 |
| Number of export partners | value | 0.71 | -0.15 | 0.08 | 0.29 | 0.66 | 0.32 | 0.51 | 0.53 |
| Foreign Direct Investment: Inward stock | percentage of world | 0.69 | -0.14 | 0.09 | 0.28 | 0.65 | 0.20 | 0.42 | 0.45 |
| Domestic Tourism Spending | usd nominal | 0.70 | -0.36 | -0.12 | 0.08 | 0.63 | 0.04 | 0.29 | 0.34 |
| Available airline seat | rank* | 0.76 | -0.22 | 0.00 | 0.20 | 0.64 | 0.11 | 0.33 | 0.35 |
| High-technology exports (current US\$) | value | 0.69 | -0.14 | 0.08 | 0.27 | 0.63 | 0.25 | 0.45 | 0.45 |
| Visitor Exports (Foreign spending) | usd real | 0.74 | -0.11 | 0.10 | 0.27 | 0.66 | 0.23 | 0.42 | 0.43 |
| No. Of Imported HS6 Digit Products | value | 0.68 | -0.17 | 0.06 | 0.27 | 0.66 | 0.29 | 0.50 | 0.51 |
| Citable documents H index | rank* | 0.71 | -0.19 | 0.06 | 0.25 | 0.67 | 0.25 | 0.47 | 0.49 |
| No. Of Exported HS6 Digit Products | value | 0.65 | -0.13 | 0.10 | 0.30 | 0.64 | 0.32 | 0.51 | 0.52 |
| GCI 4.0: Air transport | score | 0.79 | -0.07 | 0.16 | 0.35 | 0.66 | 0.28 | 0.47 | 0.46 |
| International tourism, number of arrivals | value | 0.72 | -0.16 | 0.05 | 0.24 | 0.64 | 0.18 | 0.40 | 0.43 |
| Research and development (R&D) | rank* | 0.72 | 0.01 | 0.23 | 0.39 | 0.65 | 0.36 | 0.53 | 0.51 |
| GCI 4.0: Research | rank* | 0.72 | 0.03 | 0.26 | 0.42 | 0.62 | 0.37 | 0.55 | 0.54 |
| Merchandise: Concentration and diversification indices of exports by country | diversification index* | 0.63 | -0.04 | 0.18 | 0.34 | 0.60 | 0.39 | 0.56 | 0.53 |
| Merchandise: Trade matrix by product groups, exports | value | 0.62 | -0.28 | -0.06 | 0.12 | 0.56 | 0.09 | 0.32 | 0.34 |
| Merchandise: Concentration and diversification indices of imports by country | diversification index* | 0.67 | 0.00 | 0.23 | 0.42 | 0.64 | 0.38 | 0.57 | 0.58 |

\*The correlations of the rank variables and diversification indices were multiplied by negative one.

**S2 Table.** Standalone correlations in the *infrastructure & digitalization* community. Spearman rank correlations between COVID-19 variables and indicators of the *infrastructure & digitalization* community in the worldwide dataset.

| Name | Type | dyssincefstcase | cases20dysaftfst | cases40dysaftfst | cases60dysaftfst | dyssincefstdeath | deaths20dysaftfst | deaths40dysaftfst | deaths60dysaftfst |
| --- | --- | --- | --- | --- | --- | --- | --- | --- | --- |
| Public Services | score* | 0.59 | 0.38 | 0.53 | 0.65 | 0.49 | 0.60 | 0.67 | 0.61 |
| ICT access | rank* | 0.59 | 0.38 | 0.54 | 0.65 | 0.51 | 0.63 | 0.69 | 0.62 |
| Country rank and value in the UNCTAD B2C E-commerce Index | rank* | 0.61 | 0.30 | 0.46 | 0.59 | 0.52 | 0.59 | 0.67 | 0.61 |
| 3rd pillar: Infrastructure | rank* | 0.56 | 0.33 | 0.49 | 0.62 | 0.50 | 0.60 | 0.69 | 0.64 |
| Secure Internet servers/million pop. | rank* | 0.53 | 0.38 | 0.52 | 0.64 | 0.48 | 0.63 | 0.69 | 0.65 |
| Fixed broadband subscriptions (per 100 people) | value | 0.56 | 0.30 | 0.50 | 0.61 | 0.53 | 0.61 | 0.71 | 0.66 |
| GCI 4.0: Exposure to unsafe drinking water | score* | -0.57 | -0.34 | -0.53 | -0.66 | -0.53 | -0.64 | -0.72 | -0.68 |
| GCI 4.0: 6.B Future workforce | score | 0.59 | 0.33 | 0.51 | 0.64 | 0.52 | 0.59 | 0.67 | 0.60 |
| B. Readiness subindex | value | 0.61 | 0.30 | 0.49 | 0.62 | 0.51 | 0.60 | 0.69 | 0.63 |
| B. Electricity and telephony infrastructure | rank* | 0.56 | 0.35 | 0.50 | 0.62 | 0.50 | 0.58 | 0.66 | 0.61 |
| 5th pillar Higher education and training | value | 0.61 | 0.28 | 0.45 | 0.57 | 0.60 | 0.54 | 0.64 | 0.57 |
| GDP per person employed (constant 2011 PPP \$) | value | 0.60 | 0.33 | 0.50 | 0.64 | 0.55 | 0.59 | 0.69 | 0.65 |
| Infrastructure | rank* | 0.65 | 0.24 | 0.44 | 0.59 | 0.57 | 0.52 | 0.64 | 0.60 |
| Information and communication technologies (ICTs) | score (0-100) | 0.67 | 0.19 | 0.40 | 0.56 | 0.60 | 0.50 | 0.62 | 0.58 |
| GCI 4.0: International co-inventions | score | 0.61 | 0.27 | 0.44 | 0.56 | 0.54 | 0.55 | 0.64 | 0.59 |
| GCI 4.0: Pillar 2: Infrastructure | rank* | 0.71 | 0.20 | 0.43 | 0.59 | 0.61 | 0.54 | 0.66 | 0.62 |
| 5th pillar: Skills | value | 0.58 | 0.32 | 0.47 | 0.59 | 0.51 | 0.57 | 0.63 | 0.56 |
| GNI per capita (constant 2010 US\$) | value | 0.57 | 0.33 | 0.48 | 0.64 | 0.54 | 0.60 | 0.70 | 0.66 |
| 4th pillar Health and primary education | value | 0.60 | 0.29 | 0.45 | 0.57 | 0.52 | 0.56 | 0.61 | 0.53 |
| GCI 4.0: Internet users | rank* | 0.62 | 0.33 | 0.50 | 0.63 | 0.50 | 0.60 | 0.66 | 0.61 |
| Wikipedia monthly edits | value | 0.50 | 0.34 | 0.54 | 0.67 | 0.51 | 0.69 | 0.75 | 0.71 |
| GCI 4.0: Pillar 3: ICT adoption | rank* | 0.61 | 0.30 | 0.44 | 0.56 | 0.44 | 0.53 | 0.59 | 0.53 |
| ICT PCT patents, applications/million pop. | rank* | 0.62 | 0.18 | 0.38 | 0.51 | 0.53 | 0.50 | 0.61 | 0.56 |
| GCI 4.0: Electricity infrastructure | rank* | 0.65 | 0.28 | 0.46 | 0.56 | 0.52 | 0.52 | 0.57 | 0.50 |
| Online creativity | score (0-100) | 0.51 | 0.33 | 0.50 | 0.60 | 0.46 | 0.62 | 0.70 | 0.65 |
| Researchers | rank* | 0.63 | 0.14 | 0.37 | 0.52 | 0.53 | 0.57 | 0.67 | 0.61 |
| Mortality rate, adult, female (per 1,000 female adults) | value | -0.65 | -0.35 | -0.53 | -0.65 | -0.55 | -0.60 | -0.66 | -0.61 |
| Human capital and research | score (0-100) | 0.62 | 0.18 | 0.35 | 0.50 | 0.54 | 0.49 | 0.61 | 0.56 |
| Quality of electricity supply | 1-7 best | 0.57 | 0.28 | 0.42 | 0.55 | 0.50 | 0.53 | 0.62 | 0.58 |
| Legitimacy of the State | score* | 0.61 | 0.21 | 0.40 | 0.54 | 0.50 | 0.50 | 0.61 | 0.58 |
| GCI 4.0: Mean years of schooling | rank* | 0.50 | 0.34 | 0.48 | 0.57 | 0.44 | 0.54 | 0.61 | 0.55 |
| GCI 4.0: Trademark applications | score | 0.53 | 0.36 | 0.53 | 0.63 | 0.50 | 0.60 | 0.68 | 0.64 |
| Generic top-level domains (gTLDs) | score (0-100) | 0.49 | 0.33 | 0.48 | 0.59 | 0.51 | 0.58 | 0.68 | 0.63 |
| Foreign Direct Investment: Inward stock | usd per capita | 0.42 | 0.43 | 0.54 | 0.63 | 0.37 | 0.60 | 0.63 | 0.58 |
| GCI 4.0: Skills of future workforce | rank* | 0.57 | 0.32 | 0.48 | 0.60 | 0.48 | 0.55 | 0.60 | 0.52 |
| Int'l Internet bandwidth, kb/s per user | value | 0.48 | 0.38 | 0.53 | 0.65 | 0.47 | 0.62 | 0.70 | 0.67 |
| GCI 4.0: Electrification rate | rank* | 0.57 | 0.35 | 0.53 | 0.61 | 0.50 | 0.60 | 0.66 | 0.59 |
| Secondary education gross enrollment rate, % | rank* | 0.52 | 0.31 | 0.52 | 0.65 | 0.52 | 0.59 | 0.67 | 0.63 |
| Mean years of schooling | scale (0 to 1) | 0.49 | 0.29 | 0.45 | 0.57 | 0.47 | 0.54 | 0.62 | 0.56 |
| Fixed telephone subscriptions (per 100 people) | value | 0.48 | 0.36 | 0.52 | 0.62 | 0.51 | 0.60 | 0.67 | 0.63 |
| Internet bandwidth | rank* | 0.46 | 0.43 | 0.54 | 0.62 | 0.40 | 0.62 | 0.66 | 0.61 |
| Self-employed, total (% of total employment) | percent | -0.50 | -0.35 | -0.46 | -0.60 | -0.40 | -0.56 | -0.63 | -0.60 |
| School life expectancy | rank* | 0.50 | 0.25 | 0.45 | 0.58 | 0.50 | 0.58 | 0.67 | 0.63 |
| Creative outputs | rank* | 0.59 | 0.24 | 0.42 | 0.53 | 0.51 | 0.49 | 0.60 | 0.54 |
| Environmental performance | index | 0.53 | 0.29 | 0.48 | 0.60 | 0.54 | 0.57 | 0.66 | 0.61 |
| Agriculture, value added (% of GDP) | value | -0.46 | -0.34 | -0.46 | -0.60 | -0.42 | -0.54 | -0.61 | -0.58 |
| A. Health | rank* | 0.59 | 0.30 | 0.48 | 0.60 | 0.57 | 0.55 | 0.62 | 0.58 |
| Innovation Output Sub-Index | rank* | 0.63 | 0.13 | 0.33 | 0.46 | 0.54 | 0.46 | 0.59 | 0.56 |
| GCI 4.0: Pupil-to-teacher ratio in primary education | score | 0.51 | 0.43 | 0.59 | 0.67 | 0.43 | 0.62 | 0.64 | 0.56 |
| B. Primary education | rank* | 0.54 | 0.24 | 0.37 | 0.48 | 0.41 | 0.47 | 0.51 | 0.44 |
| Health equality | scale (0 to 1) | 0.51 | 0.32 | 0.45 | 0.58 | 0.41 | 0.54 | 0.58 | 0.52 |
| Mortality rate, adult, male (per 1,000 male adults) | value | -0.64 | -0.32 | -0.49 | -0.60 | -0.58 | -0.57 | -0.64 | -0.59 |
| B. Primary education | value | 0.54 | 0.21 | 0.34 | 0.44 | 0.43 | 0.44 | 0.48 | 0.41 |
| Country-code top-level domains (ccTLDs) | score (0-100) | 0.41 | 0.31 | 0.48 | 0.58 | 0.42 | 0.57 | 0.65 | 0.60 |
| Fertility rate, total (births per woman) | value | -0.55 | -0.30 | -0.47 | -0.55 | -0.50 | -0.58 | -0.64 | -0.58 |
| Access to electricity (% of population) | value | 0.60 | 0.35 | 0.53 | 0.62 | 0.53 | 0.56 | 0.63 | 0.58 |
| Knowledge and technology outputs | rank* | 0.65 | 0.04 | 0.25 | 0.40 | 0.56 | 0.39 | 0.53 | 0.50 |
| Access to electricity, urban (% of urban population) | value | 0.65 | 0.29 | 0.47 | 0.55 | 0.52 | 0.51 | 0.58 | 0.52 |

\*The correlations of the rank and some score variables were multiplied by negative one.

**S3 Table.**
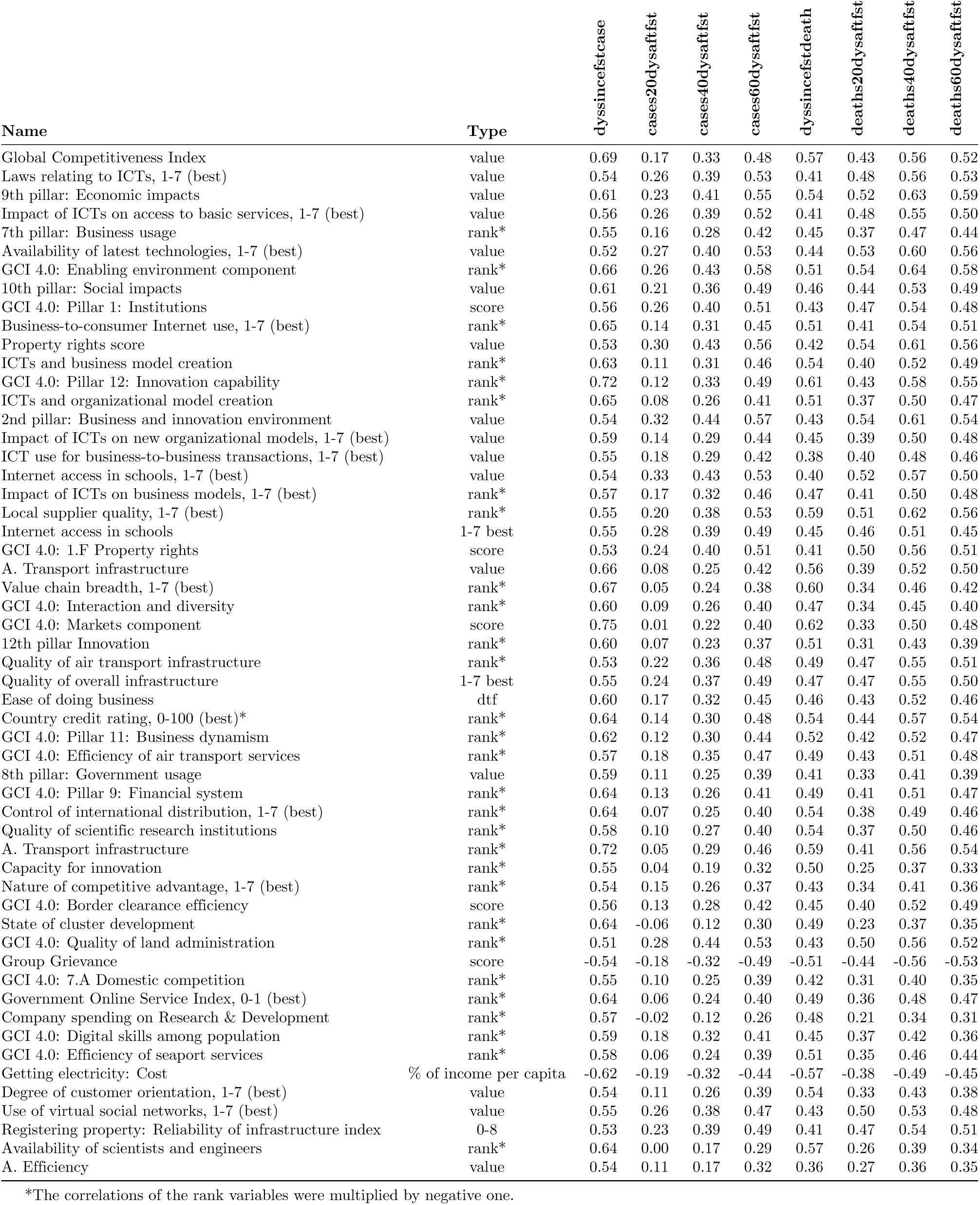
Standalone correlations in the *business & ICT* community. Spear-man rank correlations between COVID-19 variables and indicators of the *business & ICT* community in the worldwide dataset.

| Name | Type | dyssincefstcase | cases20dysaftfst | cases40dysaftfst | cases60dysaftfst | dyssincefstdeath | deaths20dysaftfst | deaths40dysaftfst | deaths60dysaftfst |
| --- | --- | --- | --- | --- | --- | --- | --- | --- | --- |
| Global Competitiveness Index | value | 0.69 | 0.17 | 0.33 | 0.48 | 0.57 | 0.43 | 0.56 | 0.52 |
| Laws relating to ICTs, 1-7 (best) | value | 0.54 | 0.26 | 0.39 | 0.53 | 0.41 | 0.48 | 0.56 | 0.53 |
| 9th pillar: Economic impacts | value | 0.61 | 0.23 | 0.41 | 0.55 | 0.54 | 0.52 | 0.63 | 0.59 |
| Impact of ICTs on access to basic services, 1-7 (best) | value | 0.56 | 0.26 | 0.39 | 0.52 | 0.41 | 0.48 | 0.55 | 0.50 |
| 7th pillar: Business usage | rank* | 0.55 | 0.16 | 0.28 | 0.42 | 0.45 | 0.37 | 0.47 | 0.44 |
| Availability of latest technologies, 1-7 (best) | value | 0.52 | 0.27 | 0.40 | 0.53 | 0.44 | 0.53 | 0.60 | 0.56 |
| GCI 4.0: Enabling environment component | rank* | 0.66 | 0.26 | 0.43 | 0.58 | 0.51 | 0.54 | 0.64 | 0.58 |
| 10th pillar: Social impacts | value | 0.61 | 0.21 | 0.36 | 0.49 | 0.46 | 0.44 | 0.53 | 0.49 |
| GCI 4.0: Pillar 1: Institutions | score | 0.56 | 0.26 | 0.40 | 0.51 | 0.43 | 0.47 | 0.54 | 0.48 |
| Business-to-consumer Internet use, 1-7 (best) | rank* | 0.65 | 0.14 | 0.31 | 0.45 | 0.51 | 0.41 | 0.54 | 0.51 |
| Property rights score | value | 0.53 | 0.30 | 0.43 | 0.56 | 0.42 | 0.54 | 0.61 | 0.56 |
| ICTs and business model creation | rank* | 0.63 | 0.11 | 0.31 | 0.46 | 0.54 | 0.40 | 0.52 | 0.49 |
| GCI 4.0: Pillar 12: Innovation capability | rank* | 0.72 | 0.12 | 0.33 | 0.49 | 0.61 | 0.43 | 0.58 | 0.55 |
| ICTs and organizational model creation | rank* | 0.65 | 0.08 | 0.26 | 0.41 | 0.51 | 0.37 | 0.50 | 0.47 |
| 2nd pillar: Business and innovation environment | value | 0.54 | 0.32 | 0.44 | 0.57 | 0.43 | 0.54 | 0.61 | 0.54 |
| Impact of ICTs on new organizational models, 1-7 (best) | value | 0.59 | 0.14 | 0.29 | 0.44 | 0.45 | 0.39 | 0.50 | 0.48 |
| ICT use for business-to-business transactions, 1-7 (best) | value | 0.55 | 0.18 | 0.29 | 0.42 | 0.38 | 0.40 | 0.48 | 0.46 |
| Internet access in schools, 1-7 (best) | value | 0.54 | 0.33 | 0.43 | 0.53 | 0.40 | 0.52 | 0.57 | 0.50 |
| Impact of ICTs on business models, 1-7 (best) | rank* | 0.57 | 0.17 | 0.32 | 0.46 | 0.47 | 0.41 | 0.50 | 0.48 |
| Local supplier quality, 1-7 (best) | rank* | 0.55 | 0.20 | 0.38 | 0.53 | 0.59 | 0.51 | 0.62 | 0.56 |
| Internet access in schools | 1-7 best | 0.55 | 0.28 | 0.39 | 0.49 | 0.45 | 0.46 | 0.51 | 0.45 |
| GCI 4.0: 1.F Property rights | score | 0.53 | 0.24 | 0.40 | 0.51 | 0.41 | 0.50 | 0.56 | 0.51 |
| A. Transport infrastructure | value | 0.66 | 0.08 | 0.25 | 0.42 | 0.56 | 0.39 | 0.52 | 0.50 |
| Value chain breadth, 1-7 (best) | rank* | 0.67 | 0.05 | 0.24 | 0.38 | 0.60 | 0.34 | 0.46 | 0.42 |
| GCI 4.0: Interaction and diversity | rank* | 0.60 | 0.09 | 0.26 | 0.40 | 0.47 | 0.34 | 0.45 | 0.40 |
| GCI 4.0: Markets component | score | 0.75 | 0.01 | 0.22 | 0.40 | 0.62 | 0.33 | 0.50 | 0.48 |
| 12th pillar Innovation | rank* | 0.60 | 0.07 | 0.23 | 0.37 | 0.51 | 0.31 | 0.43 | 0.39 |
| Quality of air transport infrastructure | rank* | 0.53 | 0.22 | 0.36 | 0.48 | 0.49 | 0.47 | 0.55 | 0.51 |
| Quality of overall infrastructure | 1-7 best | 0.55 | 0.24 | 0.37 | 0.49 | 0.47 | 0.47 | 0.55 | 0.50 |
| Ease of doing business | dtf | 0.60 | 0.17 | 0.32 | 0.45 | 0.46 | 0.43 | 0.52 | 0.46 |
| Country credit rating, 0-100 (best)* | rank* | 0.64 | 0.14 | 0.30 | 0.48 | 0.54 | 0.44 | 0.57 | 0.54 |
| GCI 4.0: Pillar 11: Business dynamism | rank* | 0.62 | 0.12 | 0.30 | 0.44 | 0.52 | 0.42 | 0.52 | 0.47 |
| GCI 4.0: Efficiency of air transport services | rank* | 0.57 | 0.18 | 0.35 | 0.47 | 0.49 | 0.43 | 0.51 | 0.48 |
| 8th pillar: Government usage | value | 0.59 | 0.11 | 0.25 | 0.39 | 0.41 | 0.33 | 0.41 | 0.39 |
| GCI 4.0: Pillar 9: Financial system | rank* | 0.64 | 0.13 | 0.26 | 0.41 | 0.49 | 0.41 | 0.51 | 0.47 |
| Control of international distribution, 1-7 (best) | rank* | 0.64 | 0.07 | 0.25 | 0.40 | 0.54 | 0.38 | 0.49 | 0.46 |
| Quality of scientific research institutions | rank* | 0.58 | 0.10 | 0.27 | 0.40 | 0.54 | 0.37 | 0.50 | 0.46 |
| A. Transport infrastructure | rank* | 0.72 | 0.05 | 0.29 | 0.46 | 0.59 | 0.41 | 0.56 | 0.54 |
| Capacity for innovation | rank* | 0.55 | 0.04 | 0.19 | 0.32 | 0.50 | 0.25 | 0.37 | 0.33 |
| Nature of competitive advantage, 1-7 (best) | rank* | 0.54 | 0.15 | 0.26 | 0.37 | 0.43 | 0.34 | 0.41 | 0.36 |
| GCI 4.0: Border clearance efficiency | score | 0.56 | 0.13 | 0.28 | 0.42 | 0.45 | 0.40 | 0.52 | 0.49 |
| State of cluster development | rank* | 0.64 | -0.06 | 0.12 | 0.30 | 0.49 | 0.23 | 0.37 | 0.35 |
| GCI 4.0: Quality of land administration | rank* | 0.51 | 0.28 | 0.44 | 0.53 | 0.43 | 0.50 | 0.56 | 0.52 |
| Group Grievance | score | -0.54 | -0.18 | -0.32 | -0.49 | -0.51 | -0.44 | -0.56 | -0.53 |
| GCI 4.0: 7.A Domestic competition | rank* | 0.55 | 0.10 | 0.25 | 0.39 | 0.42 | 0.31 | 0.40 | 0.35 |
| Government Online Service Index, 0-1 (best) | rank* | 0.64 | 0.06 | 0.24 | 0.40 | 0.49 | 0.36 | 0.48 | 0.47 |
| Company spending on Research & Development | rank* | 0.57 | -0.02 | 0.12 | 0.26 | 0.48 | 0.21 | 0.34 | 0.31 |
| GCI 4.0: Digital skills among population | rank* | 0.59 | 0.18 | 0.32 | 0.41 | 0.45 | 0.37 | 0.42 | 0.36 |
| GCI 4.0: Efficiency of seaport services | rank* | 0.58 | 0.06 | 0.24 | 0.39 | 0.51 | 0.35 | 0.46 | 0.44 |
| Getting electricity: Cost | % of income per capita | -0.62 | -0.19 | -0.32 | -0.44 | -0.57 | -0.38 | -0.49 | -0.45 |
| Degree of customer orientation, 1-7 (best) | value | 0.54 | 0.11 | 0.26 | 0.39 | 0.54 | 0.33 | 0.43 | 0.38 |
| Use of virtual social networks, 1-7 (best) | value | 0.55 | 0.26 | 0.38 | 0.47 | 0.43 | 0.50 | 0.53 | 0.48 |
| Registering property: Reliability of infrastructure index | 0-8 | 0.53 | 0.23 | 0.39 | 0.49 | 0.41 | 0.47 | 0.54 | 0.51 |
| Availability of scientists and engineers | rank* | 0.64 | 0.00 | 0.17 | 0.29 | 0.57 | 0.26 | 0.39 | 0.34 |
| A. Efficiency | value | 0.54 | 0.11 | 0.17 | 0.32 | 0.36 | 0.27 | 0.36 | 0.35 |

\*The correlations of the rank variables were multiplied by negative one.

**S4 Table.** Standalone correlations in the *regulation & corruption* community. Spearman rank correlations between COVID-19 variables and indicators of the *regulation & corruption* community in the worldwide dataset.

| Name | Type | dysincefstcase | cases20dysaftfst | cases40dysaftfst | cases60dysaftfst | dysincefstdeath | deaths20dysaftfst | deaths40dysaftfst | deaths60dysaftfst |
| --- | --- | --- | --- | --- | --- | --- | --- | --- | --- |
| Regulatory quality | rank* | 0.47 | 0.32 | 0.44 | 0.56 | 0.40 | 0.57 | 0.61 | 0.56 |
| Political environment | score (0-100) | 0.42 | 0.33 | 0.41 | 0.52 | 0.33 | 0.53 | 0.56 | 0.51 |
| Institutions | score (0-100) | 0.48 | 0.31 | 0.42 | 0.53 | 0.39 | 0.52 | 0.58 | 0.52 |
| Freedom from corruption score | value | 0.43 | 0.32 | 0.41 | 0.53 | 0.36 | 0.51 | 0.56 | 0.52 |
| Government officials in the police and the military do not use public office for private gain | total | 0.50 | 0.20 | 0.35 | 0.49 | 0.43 | 0.52 | 0.59 | 0.52 |
| Due process of law and the rights of the accused | total | 0.46 | 0.23 | 0.40 | 0.55 | 0.45 | 0.55 | 0.62 | 0.56 |
| Government regulations are applied and enforced without improper influence | total | 0.42 | 0.17 | 0.28 | 0.43 | 0.40 | 0.46 | 0.54 | 0.49 |
| The right to life and security of the person is effectively guaranteed | total | 0.32 | 0.30 | 0.47 | 0.61 | 0.38 | 0.60 | 0.65 | 0.59 |
| Corruption | scale (0 to 1)* | -0.46 | -0.21 | -0.32 | -0.47 | -0.37 | -0.48 | -0.54 | -0.49 |
| Regulatory environment | score (0-100) | 0.36 | 0.26 | 0.36 | 0.45 | 0.25 | 0.49 | 0.55 | 0.50 |
| Trading across borders : Time to import | days | -0.55 | -0.23 | -0.38 | -0.52 | -0.45 | -0.56 | -0.62 | -0.59 |
| Civil Liberties | 1 to 7 scale* | 0.22 | 0.28 | 0.40 | 0.47 | 0.34 | 0.50 | 0.54 | 0.50 |
| Freedom from arbitrary interference with privacy is effectively guaranteed | total | 0.38 | 0.19 | 0.39 | 0.54 | 0.42 | 0.52 | 0.59 | 0.54 |
| Uneven Development | score | -0.29 | -0.28 | -0.39 | -0.47 | -0.38 | -0.48 | -0.55 | -0.52 |
| Election government intimidation | scale (0 to 1)* | -0.34 | -0.25 | -0.36 | -0.43 | -0.42 | -0.47 | -0.54 | -0.49 |
| Election other voting irregularities | scale (0 to 1)* | -0.42 | -0.23 | -0.34 | -0.45 | -0.40 | -0.47 | -0.55 | -0.51 |

\*The correlations of the rank and scale variables were multiplied by negative one.

**S5 Table.** Standalone correlations in the *protectionism* community. Spear-man rank correlations between COVID-19 variables and indicators of the *protectionism* community in the worldwide dataset.

| Name | Type | dysincefstcase | cases20dysaftfst | cases40dysaftfst | cases60dysaftfst | dysincefstdeath | deaths20dysaftfst | deaths40dysaftfst | deaths60dysaftfst |
| --- | --- | --- | --- | --- | --- | --- | --- | --- | --- |
| Tariff rate, applied, simple mean, all products (%) | value | -0.28 | -0.39 | -0.48 | -0.54 | -0.29 | -0.52 | -0.55 | -0.50 |
| Tariff rate, most favored nation, weighted mean, manufactured products (%) | value | -0.40 | -0.35 | -0.45 | -0.54 | -0.35 | -0.50 | -0.56 | -0.51 |
| GCI 4.0: Trade tariffs | rank | -0.30 | -0.36 | -0.47 | -0.56 | -0.30 | -0.52 | -0.56 | -0.51 |
| Tariff rate, most favored nation, simple mean, manufactured products (%) | value | -0.36 | -0.37 | -0.47 | -0.55 | -0.32 | -0.51 | -0.56 | -0.50 |
| Tariff rate, most favored nation, weighted mean, all products (%) | value | -0.37 | -0.32 | -0.46 | -0.56 | -0.35 | -0.55 | -0.61 | -0.56 |
| Trade freedom score | value | 0.39 | 0.34 | 0.44 | 0.51 | 0.34 | 0.53 | 0.55 | 0.49 |
| Effectively applied import tariff rates on non-agricultural and non-fuel products | average | -0.34 | -0.33 | -0.42 | -0.47 | -0.36 | -0.52 | -0.56 | -0.49 |
| Share of tariff lines with international peaks, manufactured products (%) | value | -0.34 | -0.39 | -0.47 | -0.56 | -0.27 | -0.50 | -0.53 | -0.49 |

**S1 File. Metadata**. The metadata of *GovData360* and *TCdata360* indicators used.

**S2 File. Regional correlations**. Standalone correlations in the regional dataset.

**S3 File. Data generation and analysis**. Datasets were generated and analyzed with R Notebook, which can be used to update datasets and customize the analyses.

## Notes

### Competing Interest Statement

The authors have declared no competing interest.

