## Supplementary material for "The Role of Societal Aspects in the Formation of Official COVID-19 Reports: A Data-Driven Analysis": S3_File.Rmd

````
---
---

Title: The Role of Societal Aspects in the Formation of Official COVID-19 Reports: a Data-Driven Analysis

Authors: ***hidden for peer review***

Address: ***hidden for peer review***

Please cite as:
- ***hidden for peer review***

Ver. 1.0
Last update: 8 Augst, 2020
Website: ***hidden for peer review***

---
---

Workflow:

1.  Installing packages and loading libraries;
2.  Setting parameters;
3.  Generating dataset;
  3.1. Collecting GovData360 indicators;
  3.2. Collecting TCdata360 indicators;
  3.3. Collecting COVID-19 variables;
  3.4. Generating new COVID-19 variables;
  3.5. Compiling and preprocessing the joint dataset;
  3.6. Compiling the country dataset and metadata;
  3.7. Writing datasets into TSV files;
4.  Analyzing COVID-19 reports;
  4.1. Loading datasets and parameters;
  4.2. Preprocessing datasets;
  4.3. Detecting communities;
  4.4. Visualizing absolute rank correlation between COVID-19 and latent variables;
  4.5. Visualizing regional differences;
  4.6. Writing datasets into TSV files.

---

# 1. Installing packages and loading libraries

```{r,message=FALSE,warning=FALSE}

rm(list=ls(all=TRUE))

pkgs <- c("data360r","tidyr","purrr","plyr","dplyr","tidyverse","reshape2","lubridate","stringr","igraph","caret","pals","wordcloud","fmsb","tm","SnowballC")
if(length(pkgs <- setdiff(pkgs, rownames(installed.packages())))) install.packages(pkgs)
rm(pkgs)

library(data360r)
library(tidyr)
library(purrr)
library(plyr)
library(dplyr)
library(tidyverse)
library(reshape2)
library(lubridate)
library(stringr)
library(igraph)
library(caret)
library(pals)
library(wordcloud)
library(fmsb)
library(tm)
library(SnowballC)

```

# 2. Setting parameters

```{r,message=FALSE,warning=FALSE}

tw <- 20 # time window (default: 20)
ir <- 1 # increased (1) or rolled (else) time windows (default: 1)

lastyr <- 2015 # (default: 2015)
cmaxmissing <- 0.50 # (default: 0.50)
rmaxmissing <- 0.25 # (default: 0.25)

alpha <- 0.535 # (default: 0.535)
beta <- 0.828 # (default: 0.828)
gamma <- 0.770 # (default: 0.770)

par <- as.data.frame(rbind(c(lastyr,cmaxmissing,rmaxmissing,alpha,beta,gamma,tw,ir)))
colnames(par) <- c("lastyr","cmaxmissing","rmaxmissing","alpha","beta","gamma","tw","ir")

rm(list=setdiff(ls(), c("par")))

```

# 3. Generating dataset

## 3.1. Collecting GovData360 indicators

```{r,message=FALSE,warning=FALSE}

met <- get_metadata360(site = "gov", metadata_type = "indicators")
met <- met[met$periodicity=="annually",]

met[,ncol(met)+1] <- pmax(as.numeric(str_sub(met$dateRange,-4,-1))-5,as.numeric(str_sub(met$dateRange,0,4)),par$lastyr)
colnames(met)[ncol(met)] <- "first_year"
met[,ncol(met)+1] <- str_sub(met$dateRange,-4,-1)
colnames(met)[ncol(met)] <- "last_year"

met <- purrr::discard(met, is.list)
met <- met[!met$last_year=="MNaN",]
met <- met[!is.na(met$last_year),]
met <- met[!met$last_year<par$lastyr,]

cd <- as.data.frame(met$id)
cd[,2] <- met$first_year
cd[,3] <- met$last_year

colnames(cd)[1] <- "id"
colnames(cd)[2] <- "first_year"
colnames(cd)[3] <- "last_year"

temp <- get_metadata360(site = "gov", metadata_type = "countries")
rs <- as.data.frame(sort(unique(temp$iso3)))
colnames(rs)[1] <- "iso3"

k <- -1

for (i in 1:nrow(cd)){
  try(dt <- get_data360(site = "gov", indicator_id = cd[i,1], timeframes = as.numeric(cd[i,2]):as.numeric(cd[i,3]), output_type = "wide"))
  if (nrow(dt)==0) {
    try(dt <- get_data360(site = "gov", indicator_id = cd[i,1], output_type = "wide"))
    dt <- as.data.frame(dt)
    try(dt <- dt[,c(1:4,(ncol(dt)-(as.numeric(cd[i,3])-as.numeric(cd[i,2]))):ncol(dt))])
    try(colnames(dt)[(ncol(dt)-(as.numeric(cd[i,3])-as.numeric(cd[i,2]))):ncol(dt)] <- as.character(as.numeric(cd[i,2]):as.numeric(cd[i,3])))
  }
  try(yrs <- as.numeric(cd[i,3])-as.numeric(cd[i,2]))
  try(dt <- as.data.frame(dt))
  for (j in 1:yrs){
    try(dt[,ncol(dt)] <- as.data.frame(t(coalesce(t(as.numeric(dt[,ncol(dt)])),t(as.numeric(dt[,ncol(dt)-j]))))))
  }
  try(dt <- dt %>% select(1, ncol(dt)))
  try(colnames(dt)[1] <- "iso3")
  try(colnames(dt)[2] <- as.character(paste("g",cd[i,1],sep="")))
  try(rs <- join(rs,dt,by = "iso3", type = "left", match = "first"))
  rm(list=ls(pattern="dt"))
  yrs <- 1
  j <- round(i/nrow(cd),1)
  if (j > k){
    print(paste(j*100,"% of GovData360 is processed",sep=""))
  }
  k <- j
}

gov_meta <- met[,c(1:2,9,4,11,10,5,3,6)]
gov_indic <- rs
gov_meta$id <- paste("g",gov_meta$id,sep="")
ctrygeo <- temp[,c(3,2,9:10)]

rm(list=setdiff(ls(), c("ctrygeo","gov_meta","gov_indic","par")))

```

## 3.2. Collecting TCdata360 indicators;

```{r,message=FALSE,warning=FALSE}

met <- get_metadata360(site = "tc", metadata_type = "indicators")
met <- met[met$periodicity=="annually",]

met[,ncol(met)+1] <- pmax(as.numeric(str_sub(met$dateRange,-4,-1))-5,as.numeric(str_sub(met$dateRange,0,4)),par$lastyr)
colnames(met)[ncol(met)] <- "first_year"
met[,ncol(met)+1] <- str_sub(met$dateRange,-4,-1)
colnames(met)[ncol(met)] <- "last_year"

met <- purrr::discard(met, is.list)
met <- met[!met$last_year=="MNaN",]
met <- met[!is.na(met$last_year),]
met <- met[!met$last_year<par$lastyr,]

cd <- as.data.frame(met$id)
cd[,2] <- met$first_year
cd[,3] <- met$last_year

colnames(cd)[1] <- "id"
colnames(cd)[2] <- "first_year"
colnames(cd)[3] <- "last_year"

temp <- get_metadata360(site = "tc", metadata_type = "countries")
rs <- as.data.frame(sort(unique(temp$iso3)))
colnames(rs)[1] <- "iso3"

k <- -1

for (i in 1:nrow(cd)){
  try(dt <- get_data360(site = "tc", indicator_id = cd[i,1], timeframes = as.numeric(cd[i,2]):as.numeric(cd[i,3]), output_type = "wide"))
  if (nrow(dt)==0) {
    try(dt <- get_data360(site = "tc", indicator_id = cd[i,1], output_type = "wide"))
    dt <- as.data.frame(dt)
    try(dt <- dt[,c(1:4,(ncol(dt)-(as.numeric(cd[i,3])-as.numeric(cd[i,2]))):ncol(dt))])
    try(colnames(dt)[(ncol(dt)-(as.numeric(cd[i,3])-as.numeric(cd[i,2]))):ncol(dt)] <- as.character(as.numeric(cd[i,2]):as.numeric(cd[i,3])))
  }
  try(yrs <- as.numeric(cd[i,3])-as.numeric(cd[i,2]))
  try(dt <- as.data.frame(dt))
  for (j in 1:yrs){
    try(dt[,ncol(dt)] <- as.data.frame(t(coalesce(t(as.numeric(dt[,ncol(dt)])),t(as.numeric(dt[,ncol(dt)-j]))))))
  }
  try(dt <- dt %>% select(1, ncol(dt)))
  try(colnames(dt)[1] <- "iso3")
  try(colnames(dt)[2] <- as.character(paste("t",cd[i,1],sep="")))
  try(rs <- join(rs,dt,by = "iso3", type = "left", match = "first"))
  rm(list=ls(pattern = "dt"))
  yrs <- 1
  j <- round(i/nrow(cd),1)
  if (j > k){
    print(paste(j*100,"% of TCdata360 is processed",sep=""))
  }
  k <- j
}

tc_meta <- met[,c(1:2,9,4,11,10,5,3,6)]
tc_indic <- rs
tc_meta$id <- paste("t",tc_meta$id,sep="")
ctrygeo <- temp[,c(3,2,9:10)]

rm(list=setdiff(ls(), c("ctrygeo","gov_meta","gov_indic","tc_meta","tc_indic","par")))

```

## 3.3. Collecting COVID-19 variables

```{r,message=FALSE,warning=FALSE}

covd <- read.csv("https://opendata.ecdc.europa.eu/covid19/casedistribution/csv", na.strings = "", fileEncoding = "UTF-8-BOM")
test <- read_csv(url("https://raw.githubusercontent.com/owid/covid-19-data/master/public/data/testing/covid-testing-all-observations.csv"))

```

## 3.4. Generating new COVID-19 variables

```{r,message=FALSE,warning=FALSE}

case <- dcast(as.data.frame(covd),dateRep ~ countryterritoryCode, value.var = "cases", fun = sum, fill = NULL)
case <- case[order(as.Date(case$dateRep, format = "%d / %m / %Y")),]
case <- case[ , -which(names(case) %in% c("NA","N/A",""))]

death <- dcast(as.data.frame(covd),dateRep ~ countryterritoryCode, value.var = "deaths", fun = sum, fill = NULL)
death <- death[order(as.Date(death$dateRep, format = "%d / %m / %Y")),]
death <- death[ , -which(names(death) %in% c("NA","N/A",""))]

colnames(test)[7] <- "tottest"
tst <- dcast(as.data.frame(test),Date ~ `ISO code`, value.var = "tottest", fun = sum, fill = NULL)
tst <- tst[order(as.Date(tst$Date, format = "%d / %m / %Y")),]
tst[is.na(tst)] <- 0

temp <- as.data.frame(matrix(NA,ncol(case)-1,11))
temp[,1] <- colnames(case)[2:ncol(case)]

tmp <- as.data.frame(matrix(NA,ncol(tst)-1,6))
tmp[,1] <- colnames(tst)[2:ncol(tst)]

colnames(temp)[1] <- "countryterritoryCode"
colnames(temp)[2] <- "dyssincefstcase"
colnames(temp)[3] <- paste("cases",par$tw,"dysaftfst",sep="")
colnames(temp)[4] <- paste("cases",par$tw*2,"dysaftfst",sep="")
colnames(temp)[5] <- paste("cases",par$tw*3,"dysaftfst",sep="")
colnames(temp)[6] <- paste("cases",par$tw*4,"dysaftfst",sep="")
colnames(temp)[7] <- "dyssincefstdeath"
colnames(temp)[8] <- paste("deaths",par$tw,"dysaftfst",sep="")
colnames(temp)[9] <- paste("deaths",par$tw*2,"dysaftfst",sep="")
colnames(temp)[10] <- paste("deaths",par$tw*3,"dysaftfst",sep="")
colnames(temp)[11] <- paste("deaths",par$tw*4,"dysaftfst",sep="")
colnames(tmp)[1] <- "countryterritoryCode"
colnames(tmp)[2] <- "dyssincefsttest"
colnames(tmp)[3] <- paste("tests",par$tw,"dysaftfst",sep="")
colnames(tmp)[4] <- paste("tests",par$tw*2,"dysaftfst",sep="")
colnames(tmp)[5] <- paste("tests",par$tw*3,"dysaftfst",sep="")
colnames(tmp)[6] <- paste("tests",par$tw*4,"dysaftfst",sep="")

for (i in 2:ncol(case)){
  csum <- cumsum(case[,i])
  n <- min(which(csum != 0))
  temp[i-1,2] <- length(csum[!csum==0])
  if (ymd(as.Date(case[n,1],format = "%d / %m / %Y"))+(par$tw-1) <= max((as.Date(case[,1],format = "%d / %m / %Y")),na.rm = TRUE)) {
    temp[i-1,3] <- sum(case[n:(n+(par$tw-1)),i],na.rm=TRUE)
  }
  if (ymd(as.Date(case[n,1],format = "%d / %m / %Y"))+(par$tw*2-1) <= max((as.Date(case[,1],format = "%d / %m / %Y")),na.rm = TRUE)) {
    temp[i-1,4] <- sum(case[n:(n+(par$tw*2-1)),i],na.rm=TRUE)
  }
  if (ymd(as.Date(case[n,1],format = "%d / %m / %Y"))+(par$tw*3-1) <= max((as.Date(case[,1],format = "%d / %m / %Y")),na.rm = TRUE)) {
    temp[i-1,5] <- sum(case[n:(n+(par$tw*3-1)),i],na.rm=TRUE)
  }
  if (ymd(as.Date(case[n,1],format = "%d / %m / %Y"))+(par$tw*4-1) <= max((as.Date(case[,1],format = "%d / %m / %Y")),na.rm = TRUE)) {
    temp[i-1,6] <- sum(case[n:(n+(par$tw*4-1)),i],na.rm=TRUE)
  }
}

for (i in 2:ncol(death)){
  csum <- cumsum(death[,i])
  n <- min(which(csum != 0))
  temp[i-1,7] <- length(csum[!csum==0])
  if (sum(csum)>0){
    if (ymd(as.Date(death[n,1],format = "%d / %m / %Y"))+(par$tw-1) <= max((as.Date(death[,1],format = "%d / %m / %Y")),na.rm = TRUE)) {
      temp[i-1,8] <- sum(death[n:(n+(par$tw-1)),i],na.rm=TRUE)
    }
    if (ymd(as.Date(death[n,1],format = "%d / %m / %Y"))+(par$tw*2-1) <= max((as.Date(death[,1],format = "%d / %m / %Y")),na.rm = TRUE)) {
      temp[i-1,9] <- sum(death[n:(n+(par$tw*2-1)),i],na.rm=TRUE)
    }
    if (ymd(as.Date(death[n,1],format = "%d / %m / %Y"))+(par$tw*3-1) <= max((as.Date(death[,1],format = "%d / %m / %Y")),na.rm = TRUE)) {
      temp[i-1,10] <- sum(death[n:(n+(par$tw*3-1)),i],na.rm=TRUE)
    }
    if (ymd(as.Date(death[n,1],format = "%d / %m / %Y"))+(par$tw*4-1) <= max((as.Date(death[,1],format = "%d / %m / %Y")),na.rm = TRUE)) {
      temp[i-1,11] <- sum(death[n:(n+(par$tw*4-1)),i],na.rm=TRUE)
    }
  }else{
    temp[i-1,8] <- 0
    temp[i-1,9] <- 0
    temp[i-1,10] <- 0
    temp[i-1,11] <- 0
  }
}

for (i in 2:ncol(tst)){
  csum <- (tst[,i])
  n <- min(which(csum != 0))
  tmp[i-1,2] <- length(csum[!csum==0])
  if (is.finite(n)){
    if (ymd(as.Date(tst[n,1],format = "%d / %m / %Y"))+(par$tw-1) <= max((as.Date(tst[,1],format = "%d / %m / %Y")),na.rm = TRUE)) {
      tmp[i-1,3] <- max(tst[n:(n+(par$tw-1)),i],na.rm=TRUE)-tst[n,i]
    }
    if (ymd(as.Date(tst[n,1],format = "%d / %m / %Y"))+(par$tw*2-1) <= max((as.Date(tst[,1],format = "%d / %m / %Y")),na.rm = TRUE)) {
      tmp[i-1,4] <- max(tst[n:(n+(par$tw*2-1)),i],na.rm=TRUE)-tst[n,i]
    }
    if (ymd(as.Date(tst[n,1],format = "%d / %m / %Y"))+(par$tw*3-1) <= max((as.Date(tst[,1],format = "%d / %m / %Y")),na.rm = TRUE)) {
      tmp[i-1,5] <- max(tst[n:(n+(par$tw*3-1)),i],na.rm=TRUE)-tst[n,i]
    }
    if (ymd(as.Date(tst[n,1],format = "%d / %m / %Y"))+(par$tw*4-1) <= max((as.Date(tst[,1],format = "%d / %m / %Y")),na.rm = TRUE)) {
      tmp[i-1,6] <- max(tst[n:(n+(par$tw*4-1)),i],na.rm=TRUE)-tst[n,i]
    }
  }
}

temp <- join(temp,tmp,by = "countryterritoryCode", type = "left", match = "first")
tmp <- covd %>% dplyr::group_by(countryterritoryCode) %>% dplyr::summarise(totcase = sum(cases,na.rm=TRUE), totdeath = sum(deaths,na.rm=TRUE) ,population = first(popData2019))
temp <- join(temp,tmp,by = "countryterritoryCode", type = "left", match = "first")
colnames(test)[2] <- "countryterritoryCode"
tmp <- test %>% dplyr::group_by(countryterritoryCode) %>% dplyr::summarise(tottest = max(tottest,na.rm=TRUE))
covid <- join(temp,tmp,by = "countryterritoryCode", type = "left", match = "first")
colnames(covid)[1] <- "iso3"

rm(list=setdiff(ls(), c("covid","ctrygeo","gov_indic","tc_indic","gov_meta","tc_meta","par")))

```

## 3.5. Compiling and preprocessing the joint dataset

```{r,message=FALSE,warning=FALSE}

temp <- join(gov_indic,tc_indic,by = "iso3", type = "left", match = "first")
temp <- temp[, which(colMeans(!is.na(temp)) > (1-par$cmaxmissing))]
temp <- temp[which(rowMeans(!is.na(temp)) > (1-par$rmaxmissing)),]
cty <- as.data.frame(temp[,1])
tmp <- temp[2:ncol(temp)]
duplicated.columns <- duplicated(t(tmp))
tmp <- tmp[, !duplicated.columns]
temp <- as.data.frame(cbind(cty,tmp))
colnames(temp)[1] <- "iso3"

joined <- join(covid,temp,by = "iso3", type = "right", match = "first")
joined <- joined[!joined$iso3=="NA",]
pop <- joined$population
ctrygeo <- join(ctrygeo,joined[,c(1,19)],by = "iso3", type = "left", match = "first")

joined <- joined[,-19]

joined[,c(3:6,8:11,13:19)] <- joined[,c(3:6,8:11,13:19)]/pop
joined <- joined[ , -which(names(joined) %in% c("g31442","g40942","t352","t3295"))]

rm(temp,tmp,cty,duplicated.columns)

```

## 3.6. Compiling the country dataset and metadata

```{r,message=FALSE,warning=FALSE}

meta <- rbind(gov_meta,tc_meta)
meta <- meta[meta$id %in% colnames(joined)[20:ncol(joined)],]
ctrygeo <- ctrygeo[ctrygeo$iso3 %in% joined$iso3,]
meta$definition <- str_replace_all(meta$definition, "[\r\n]" , "")

rm(gov_indic, tc_indic, gov_meta, tc_meta)

```

## 3.7 Writing datasets into TSV files

```{r,message=FALSE,warning=FALSE}

write.table(ctrygeo, "country_data.txt", row.names=FALSE, sep="\t",quote=FALSE)
write.table(meta, "metadata.txt", row.names=FALSE, sep="\t",quote=FALSE)
write.table(joined, "joint_dataset.txt", row.names=FALSE, sep="\t",quote=FALSE)
write.table(par, "parameters.txt", row.names=FALSE, sep="\t",quote=FALSE)

```

# 4. Analyzing COVID-19 reports

## 4.1. Loading datasets and parameters

```{r,message=FALSE,warning=FALSE}

try(data <- read.csv('joint_dataset.txt', header = TRUE, sep="\t", dec = ".",stringsAsFactors = TRUE))
try(feat <- read.csv('metadata.txt', header = TRUE, sep="\t", dec = ".",stringsAsFactors = FALSE))
try(par <- read.csv('parameters.txt', header = TRUE, sep="\t",stringsAsFactors = FALSE))

needed <- c("dyssincefstcase",paste("cases",par$tw,"dysaftfst",sep=""),paste("cases",par$tw*2,"dysaftfst",sep=""),paste("cases",par$tw*3,"dysaftfst",sep=""),"dyssincefstdeath",
          paste("deaths",par$tw,"dysaftfst",sep=""),paste("deaths",par$tw*2,"dysaftfst",sep=""),paste("deaths",par$tw*3,"dysaftfst",sep=""))

```

## 4.2. Preprocessing datasets

```{r,message=FALSE,warning=FALSE}

X <- data[,20:length(colnames(data))]
X <- scale(data.matrix(X, rownames.force = NA))
rem <- data.frame(id=colnames(X),num=seq(1,ncol(X)))
removex <- feat$id[feat$valueType=="StdErr" | feat$valueType=="NumSrc"]
remove_cols <- rem[is.element(rem$id,removex),]
X <- X[,-remove_cols$num]

nzv <- nearZeroVar(X)
X <- X[, -nzv]
tmp <- cor(X,method="spearman",use="pairwise.complete.obs")
tmp[is.na(tmp)] <- 0
diag(tmp) <- 0
hc <- findCorrelation(tmp, cutoff = 0.95)
X <- X[,-c(hc)]

y <- data[,2:19]
y <- data.matrix(y, rownames.force = NA)
y <- y[,needed]

Xnames <- colnames(X)
ynames <- colnames(y)
X <- as.data.frame(X)
y <- as.data.frame(y)

if (par$ir!=1){
 y[,4] <- y[,4] - y[,3]
 y[,3] <- y[,3] - y[,2]
 y[,8] <- y[,8] - y[,7]
 y[,7] <- y[,7] - y[,6]
}

df <- cbind(X,y)
ny <- dim(y)[2]

```

## 4.3. Detecting communities

```{r,message=FALSE,warning=FALSE}

cormat0 <- cor(df,method="spearman",use="pairwise.complete.obs")
rownames(cormat0) <- c(Xnames,ynames)
colnames(cormat0) <- c(Xnames,ynames)
cormat <- abs(cormat0)
cormat[is.na(cormat)] <- 0

thold <- par$alpha

tholdvec <- apply(cormat[(dim(cormat)[1]-ny+1):dim(cormat)[1],],2,max)>thold
mat <- cormat[tholdvec,tholdvec]

thold <- par$beta
pclim <- par$gamma

xmat <- mat[-c((dim(mat)[1]-ny+1):dim(mat)[1]),-c((dim(mat)[1]-ny+1):dim(mat)[1])]
mat2 <- ifelse(xmat<thold,0,xmat)
diag(mat2) <- 0
g <- graph_from_adjacency_matrix(mat2,mode = c("upper"), weighted = TRUE)
Isolated <- which(degree(g)==0)
g2 <- delete.vertices(g, Isolated)
cl <- cluster_louvain(g,weights = E(g)$weight)
memb <- membership(cl)

cl_rank <- as.data.frame(table(cl$membership))
colnames(cl_rank) <- c("member1","Freq")

ycor <- apply(cormat[(dim(cormat)[1]-ny+1):dim(cormat)[1],],2,max)[tholdvec]
ycor <- ycor[1:(length(ycor)-ny)]
ycordf <- data.frame(m=as.factor(memb),ycor=ycor)
meancor <- as.data.frame(aggregate(ycordf$ycor, list(ycordf$m), mean))
cl_rank$ycor <- meancor$x

cl_rank <- cl_rank[cl_rank$Freq>6,]
cl_rank <- cl_rank[order(cl_rank$ycor,decreasing = TRUE),][1:10,]
cl_rank$rank <- seq(1,length(cl_rank$member1))
col <- cols25(25)
col <- c("blue","red","purple","#0fd61c","#fac832","cyan")
cols <- rep("white",length(memb))

for (i in 1:length(memb)){
  if (is.element(memb[i],cl_rank$member1)==TRUE){
    cols[i] <- col[cl_rank$rank[cl_rank$member1==memb[i]]]
  }
}

rescale = function(x,a,b,c,d){c + (x-a)/(b-a)*(d-c)}
centrality_df<-data.frame(id=V(g)$name)

for (i in 1:length(unique(memb))){
  g2 <- induced_subgraph(g,memb==unique(memb)[i])
  ec <- eigen_centrality(g2)
  b <- ec$vector  
  b[is.nan(b)] <- 0
  br <- rescale(b, min(b), max(b), 0, 1)
  for (j in 1:length(b)){
    centrality_df$centrality[which(as.character(centrality_df$id)==names(b)[j])] <- b[j]
    centrality_df$centrality_res[which(as.character(centrality_df$id)==names(br)[j])] <- br[j]
  }
}

centrality_df$centrality_res[is.nan(centrality_df$centrality_res)] <- 0
centrality_df$size_rescaled <- rescale(centrality_df$centrality_res, min(centrality_df$centrality_res), max(centrality_df$centrality_res), 3, 10)

feat_df <- data.frame(id=V(g)$name)
feat_df$idnum <- V(g)
feat_df <- merge(feat_df,feat,sort=F,all.x=T)
feat_df$community <- memb
feat_df$centrality_comm <- centrality_df$centrality
feat_df$filter <- is.element(feat_df$community,cl_rank$member1)
feat_df <- feat_df[feat_df$filter==TRUE,]
for(i in 1:nrow(feat_df)){feat_df$rank[i] <- cl_rank$rank[cl_rank$member1==feat_df$community[i]]}
feat_df <- feat_df[order(feat_df$rank,-feat_df$centrality_comm),]

comms <- sum(!is.na(cl_rank$Freq))
sumcormat <- matrix(rep(0,ny*comms),nrow=ny,ncol=comms)
datamat <- matrix(rep(0,nrow(data)*comms),nrow=nrow(data),ncol=comms)
remall <- vector()

for (j in 1:comms){
  limit <- pclim
  remvec <- vector()
  avgcor <- vector()
  featcnt <- vector()
  subvec0 <- V(g)$name[memb==cl_rank$member1[cl_rank$rank==j]]
  for (i in 1:length(subvec0)){
    subvec <- subvec0[is.element(subvec0,remvec)==FALSE]
    try(g2 <- induced_subgraph(g,subvec))
    ec <- eigen_centrality(g2)
    vardf <- df[,names(ec$vector)]
    vardf <- as.data.frame(ifelse((cor(vardf,method="spearman",use="pairwise.complete.obs")[which.max(ec$vector),]<0)==TRUE,vardf*-1,vardf))
    vardf[is.na(vardf)] <- 0
    vardf <- as.data.frame(vardf)
    b <- rowSums(ec$vector/sum(ec$vector)*vardf)
    vardf$pc <- b
    if (min(abs(cor(vardf,method="spearman",use="pairwise.complete.obs")[,"pc"]))>limit){
      break
    }
    rem <- which.min(abs(cor(vardf,method="spearman",use="pairwise.complete.obs")[,"pc"]))
    remvec <- c(remvec,names(rem))
    avgcor <- c(avgcor,min(abs(cor(vardf,method="spearman",use="pairwise.complete.obs")[,"pc"])))
    featcnt <- c(avgcor,mean(abs(cor(vardf,method="spearman",use="pairwise.complete.obs")[,"pc"])))
  }
  sumcormat[,j] <- cor(cbind(y,vardf$pc),method="spearman",use="pairwise.complete.obs")[1:ny,"vardf$pc"] 
  rownames(sumcormat) <- needed  
  datamat[,j] <- b
  remall <- c(remall,remvec)
  print(j)
}  

corrdata <- cbind(y,datamat)
cmat <- cor(corrdata,method="spearman",use="pairwise.complete.obs")
feat_dfr <- feat_df[is.element(feat_df$id,remall)==FALSE,]
absmax <- function(x) { x[which.max( abs(x) )]}
amaxvec <- apply(cormat0[feat_dfr$id,needed],1,absmax)
maxloc <- apply(cormat[feat_dfr$id,needed],1,which.max)

for (i in 1: length(V(g)$name)){
  if (is.element(V(g)$name[i],remall)==TRUE){
    cols[i] <- "darkgrey"
  }
}

V(g)$color <- cols
ycor <- apply(cormat[(dim(cormat)[1]-ny+1):dim(cormat)[1],],2,max)[tholdvec]
ycor2 <- rescale(ycor, min(ycor), max(ycor), 4, 11)
V(g)$size <- ycor2[1:(length(ycor2)-ny)]
Isolated <- which(degree(g)==0)
g2 <- delete.vertices(g, Isolated)
plot(g2,vertex.label=NA,edge.color=rgb(0.4,0.4,0.4,alpha=0.5))
par(mfrow=c(2,4))
par(mar=c(0,0,0,0))

for (i in 1:comms){
  text <- feat_dfr$name[feat_dfr$rank==i]
  remove_vec <- c("capita","per","value","constant","using","usage","rank","gci","best","current","index","pillar","quality","score","due","use","relating","subindex","index","mean","service","services","access","contribution","gain","average","total","rate","country","data","world","number")
  
  docs <- Corpus(VectorSource(text))
  docs <- tm_map(docs, content_transformer(tolower))
  docs <- tm_map(docs, removeNumbers)
  docs <- tm_map(docs, removeWords, stopwords("english"))
  docs <- tm_map(docs, removeWords, c("blabla1", "blabla2")) 
  docs <- tm_map(docs, removePunctuation)
  docs <- tm_map(docs, stripWhitespace)
  
  dtm <- TermDocumentMatrix(docs)
  m <- as.matrix(dtm)
  v <- sort(rowSums(m),decreasing=TRUE)
  d <- data.frame(word = names(v),freq=v)
  d$filter <- is.element(d$word,remove_vec)
  d <- d[d$filter==FALSE,]
  try(d$rank <- 1:nrow(d))
  d$color <- as.character(ifelse(d$rank<nrow(d)*0.1,col[i],"black"))
  wcolor <- rep("black",nrow(d))
  wcolor[1:(round(length(d$freq)*0.2,0)+1)] <- col[i]
  try(wordcloud(words = d$word, freq = d$freq, min.freq = 1, max.words=50, random.order=FALSE, rot.per=0.35, colors=as.character(wcolor),ordered.colors=TRUE))
}

```

## 4.4. Visualizing absolute rank correlation between COVID-19 and latent variables

```{r,message=FALSE,warning=FALSE}

# Define the number of dimensions of the radar charts (the number of topics to visualize).
num_dimensions <- 5

# Define the name of the topics.
covtopics <- c("Tourism & Trade","Infrastructure","Business","Transparency","Protectionalism")

# Define the desired grouping of target variables by setting the list below. A radar chart will be generated for each element of the list. Note that, only the elements of variable "needed" can be selected into "radar_list".
radar_list <- list(
                    Appereance=c("dyssincefstcase","dyssincefstdeath"),
                    Cases=c(paste("cases",par$tw,"dysaftfst",sep=""),paste("cases",par$tw*2,"dysaftfst",sep=""),paste("cases",par$tw*3,"dysaftfst",sep="")),
                    Deaths=c(paste("deaths",par$tw,"dysaftfst",sep=""),paste("deaths",par$tw*2,"dysaftfst",sep=""),paste("deaths",par$tw*3,"dysaftfst",sep=""))
                  )
                   
ceilf <- function(x, l=1) round(x + 5*10^(-l-1), l)
lims <- as.data.frame(matrix(rep(ceilf(max(abs(sumcormat)),1), num_dimensions*2), nrow=2, ncol=num_dimensions))
lims[2,] <- 0
radardata <- rbind(lims,abs(sumcormat[,1:num_dimensions]))
colnames(radardata) <- covtopics[1:num_dimensions]
rectangles <- c("#a6c2f7","#0554e8","black")
w <- 3
plty <- c(1,1,1,1)

for (i in 1:length(radar_list)){
  par(xpd=TRUE)
  radarchart(radardata[c(1,2,radar_list[[i]]),],axistype=2,centerzero=TRUE,cglcol="gray84",axislabcol="black",palcex=1.3,vlcex=1.3,cglty=2,pcol=rectangles,plty=plty,pty=32,plwd=w,title=names(radar_list[i]))
  legend("topright", inset=c(-0.05,0),
         legend=radar_list[[i]],
         col=rectangles[1:length(radar_list[[i]])],
         lty=plty[1:length(radar_list[[i]])],
         lwd=3,
         bty="n")
}

```

## 4.5. Visualizing regional differences

```{r,message=FALSE,warning=FALSE}

# Define the number of dimensions of the radar charts (the number of topics to visualize).
num_dimensions <- 5
num_dimensions <- min(comms,num_dimensions)

# Define the color of the radar lines. The order is the following: Europe, Asia, Americas, Africa.
rectangles <- c("blue","red","gold","green")

reg <- read_csv(url("https://raw.githubusercontent.com/lukes/ISO-3166-Countries-with-Regional-Codes/master/all/all.csv"))
codes <- data.frame(iso3=data[,1])
regions <- merge(codes,reg[,c("alpha-3","region")],by.x="iso3",by.y="alpha-3",all.x=TRUE,all.y=FALSE)
region_data <- cbind(regions$region, y, datamat)
cont <- c("Europe","Asia","Americas","Africa")
cordf_all <- as.data.frame(matrix(rep(0,num_dimensions+2),nrow=1,ncol=num_dimensions+2))
colnames(cordf_all) <- c(covtopics[1:num_dimensions],"variable","region")

for (i in 1:4){
  filtered0 <- region_data[region_data$region==cont[i],2:ncol(region_data)]
  filtered <- data.matrix(filtered0, rownames.force = NA)
  cormat <- cor(filtered,method="spearman",use="pairwise.complete.obs")
  cormat <- abs(cormat[1:ny,(ny+1):(ny+num_dimensions)])
  colnames(cormat) <- covtopics[1:num_dimensions]
  cordf <- as.data.frame(cormat)
  cordf$variable <- as.character(rownames(cordf))
  cordf$region <- cont[i]
  cordf_all <- rbind(cordf_all,cordf)
}

for (i in 1:ny){
  radardata <- cordf_all[cordf_all$variable==needed[i],]
  rownames(radardata) <- radardata$region
  radardata <- radardata[,1:num_dimensions]
  lims <- as.data.frame(matrix(rep(ceilf(max(abs(sumcormat)),1),num_dimensions*2),nrow=2,ncol=num_dimensions))
  lims[2,] <- 0
  colnames(lims) <- colnames(radardata)
  radardata <- rbind(lims,radardata)
  radarchart(radardata,axistype=2,cex.main=1.5,cglcol="gray84",cglty=2,centerzero=TRUE,pcol=rectangles,axislabcol="black",plty=1,pty=32,plwd=2,title=needed[i],vlcex=1.3,palcex=1.3)
  legend("topright", inset=c(-0.01,0),legend=cont,col=rectangles,lty=1,bty="n",lwd=3)
}

```

## 4.6 Writing datasets into TSV files

```{r,message=FALSE,warning=FALSE}

regional_correlations <- data.frame()

for (i in 1:4){
  df2 <- df[regions$region==cont[i],]
  cor_cont <- cor(df2,method="spearman",use="pairwise.complete.obs")
  contdf <- as.data.frame(cor_cont[as.character(feat_dfr$id),ynames])
  contdf$id <- feat_dfr$id
  contdf$name <- feat_dfr$name
  contdf$community <- feat_dfr$rank
  contdf$type <- feat_dfr$valueType
  contdf$region <- cont[i]
  regional_correlations <- rbind(regional_correlations,contdf)
  row.names(regional_correlations) <- NULL
  print(paste(cont[i]," data collected.",sep=""))
}
write.table(regional_correlations, "regional_correlations.txt", row.names=FALSE, sep="\t",quote=FALSE)
print("Correlations calculated region-wise exported as regional_correlations.txt.")

correlations_all <- as.data.frame(cormat0[as.character(feat_dfr$id),ynames])
correlations_all$id <- feat_dfr$id
correlations_all$name <- feat_dfr$name
correlations_all$community <- feat_dfr$rank
correlations_all$type <- feat_dfr$valueType
write.table(correlations_all, "correlations_all.txt", row.names=FALSE, sep="\t",quote=FALSE)
print("Correlations without regional subsetting exported as correlations_all.txt.")

```
````
